# Prevalence of motor neuron disease and its clinical subtypes in the Valencian Community, Spain: a population-based cross-sectional study using linked electronic health records

**DOI:** 10.64898/2026.10.07.26364981

**Authors:** Pablo Ros-Arlanzón, Juan Francisco Vázquez-Costa, Ana Merchán-Cabanillas, Rosa Vilar-Ventura, Ana Pareja, Laura Lorente-Gómez, Elena Ronda-Pérez, Carmen Díaz-Marín

## Abstract

**Background:** To determine the crude and age-adjusted point prevalence of motor neuron disease (MND) and its phenotypes (classical amyotrophic lateral sclerosis [ALS], primary lateral sclerosis [PLS] and progressive muscular atrophy [PMA]) using a comprehensive multi-source case ascertainment strategy.

**Methods:** Cross-sectional population-based study with a prevalence date of 1 January 2025. Cases were identified through five complementary regional data sources and confirmed by clinical review against the revised El Escorial and Gold Coast criteria. Cases were classified by phenotype as ALS, PLS or PMA. The reference population was the population aged 15 years or older in Valencian Community (n=4,433,999). Crude prevalence was calculated per 100,000 with 95% Poisson confidence intervals; age-adjusted rates were obtained by direct standardisation to the 2013 European Standard Population.

**Results:** A total of 374 confirmed MND cases were identified. The overall crude prevalence of MND was 8.43 per 100,000 (95% CI: 7.60–9.29) and the age-adjusted prevalence was 7.85 per 100,000 (95% CI: 7.05–8.65). By phenotype, ALS accounted for 325 cases (86.9%; crude prevalence 7.33 per 100,000), PLS for 26 (7.0%; 0.59 per 100,000) and PMA for 23 (6.1%; 0.52 per 100,000). Prevalence was consistent across provinces. Mean age was 65.9 years, the male-to-female ratio was 1.37:1, and 83.4% were receiving riluzole.

**Conclusions:** This is the first comprehensive, multi-source population-based estimate of MND prevalence and its subtypes in the Valencian Community. The estimate is higher than the previous single-unit figure and consistent with recent European data, underscoring the value of multi-source ascertainment.

## 1. Introduction

Motor neuron diseases (MND) are a group of progressive neurodegenerative disorders characterised by the degeneration of upper and/or lower motor neurons (UMN and LMN) harbouring intracellular deposits of misfolded proteins (usually TDP-43). Amyotrophic lateral sclerosis (ALS) is the most frequent phenotype, comprising approximately 85–90% of cases, and affecting both UMN and LMN. Less common phenotypes include those patients with restricted degeneration of UMN (named primary lateral sclerosis, PLS) or LMN (progressive muscular atrophy, PMA) [1,2]. Although these subtypes share pathophysiological mechanisms, their demographical and clinical differences have resulted in different diagnostic codes.[2]

Accurate, population-based epidemiological data are essential for health-service planning and clinical-trial design. Recent studies have reported ALS prevalence figures of between 4.3 and 6.0 per 100,000, with marked geographical heterogeneity,[3–5] whereas population-based estimates for PLS and PMA are exceedingly rare.

In Spain, a previous study estimated the prevalence of MND and its subtypes in Catalonia and Valencia using data from a single referral unit in each region.[6] For Valencia, that study estimated an ALS prevalence of 3.25 per 100,000 (2013–2018), and noted that the dispersed geography of the Valencian population could limit access to the single unit and affect case capture.[6]

Since then, the Valencian Community has consolidated a network of four MND reference units covering its entire territory, with integrated electronic health information systems linking primary care, specialised care, pharmacy and hospital data. The aim of the present study was to estimate the crude and age-adjusted point prevalence of MND and of its subtypes (ALS, PLS and PMA) as of 1 January 2025 using a multi-source approach.

## 2. Material and methods

### 2.1. Study design and setting

Observational, descriptive, population-based cross-sectional study estimating the point prevalence of MND in the Valencian Community (eastern Spain, ~5 million inhabitants). The reference date was 1 January 2025. Healthcare is universal, and every resident is automatically assigned an individual health card and electronic health record (EHR), ensuring near-complete coverage. The region is served by four MND reference units, which receive referrals from other hospitals: Hospital General Universitario Dr. Balmis (Alicante), Hospital Universitari i Politècnic La Fe and Hospital Clínico Universitario (Valencia), and Hospital General Universitario de Castellón.

### 2.2. Study population

All residents with a confirmed diagnosis of MND alive on the reference date. The denominator was the registered population aged 15 years or older at 1 January 2025, obtained from the Primary Care Information System (SIAP) [7] (n=4,433,999).

### 2.3. Case identification

Potential cases were identified through five complementary sources: (1) ICD-10 (G12.2, G12.20– G12.29) and ICD-9 (335.2x) codes in the EHR (diagnosis related to these codes are shown in Supplementary Table S1); (2) riluzole prescription and dispensing records; (3) the hospital discharge dataset; and (4) the internal registries of each reference unit; and (5) PROSIGA, the regional outpatient care information system, which provides access to primary and specialised care clinical records for research purposes.

### 2.4. Case confirmation and phenotyping

All identified patients underwent systematic clinical review by experienced neurologists of the corresponding reference unit. Confirmed ALS was defined by meeting either the revised El Escorial [8] or Gold Coast criteria [9] at the reference date and showing a clinical course suggestive of classical ALS. PLS cases were those meeting clinical criteria for definite PLS at the reference date and showing a clinical course suggestive of PLS.[10] PMA cases were those patients showing clinical or neurophysiological signs of LMN impairment in at least 2 regions at the reference date, together with the absence of UMN signs 4 or more years from symptom onset, and showing a clinical course suggestive of PMA (i.e. spinal onset with lack of respiratory insufficiency or significant bulbar impairment during the first 4 years of disease).[11] Well-established ALS variants (including bulbar- and spinal-onset, flail leg, ALS–frontotemporal dementia or ALS–parkinsonism) were grouped under ALS. Patients initially identified as cases but with alternative diagnoses after careful review of the medical record and appropriate diagnostic work-up were excluded. Age at the reference date and sex were collected.

### 2.5. Genetic testing

Genetic results were collected when available. Genetic testing was performed as part of routine clinical care at the reference laboratories of the Valencian Community, and its extent varied according to the date of diagnosis, the referring unit and the clinical indication. *C9orf72* hexanucleotide repeat expansion was analysed separately by a specific assay (repeat-primed PCR) that quantifies the number of repeats; alleles were considered pathological at or above the established expansion threshold. Sequencing of other ALS-related genes was performed either as targeted single-gene analysis (most often SOD1) or through a multigene panel that included, in addition to core ALS/FTD genes, genes associated with ALS mimics (*ALS2, ANG, ANXA11, ATXN2, BSCL2, CFAP410 (C21orf2), CHCHD10, CHMP2B, DAO, DCTN1, EPHA4, ERBB4, FIG4, FUS, GLE1, HNRNPA1, HNRNPA2B1, KIF5A, MATR3, NEFH, NEK1, NIPA1, OPTN, PARK7 (DJ-1), PFN1, PRPH, SETX, SIGMAR1, SOD1, SPAST, SPG11, SPG7, SQSTM1, TAF15, TARDBP, TBK1, TIA1, TUBA4A, UBQLN2, UNC13A, VAPB* and *VCP*). Consequently, not all patients were tested for the same set of genes, and the denominator of tested patients differs by gene. Variants were interpreted following standard ACMG criteria and classified as pathogenic, likely pathogenic or of uncertain significance.[12]

### 2.6. Statistical analysis

Crude prevalence was calculated per 100,000 with 95% Poisson confidence intervals, overall and by phenotype and province and/or health department. Age-adjusted prevalence was obtained by direct standardisation to the 2013 European Standard Population(13); age-specific standard weights were restricted to the population aged 15 years and older, consistent with the study denominator, and renormalised to sum to one; 95% confidence intervals for the standardised rates were derived from the variance of the weighted sum of age-specific rates using a normal approximation.

### 2.7. Ethics approval

The study was approved by the Committee of Ethics of Clinical Research of the Dr. Balmis University Hospital (reference PI2026-018). Given the retrospective, registry-based design, individual informed consent was waived.

## 3. Results

### 3.1. Case ascertainment

After pooling the five sources and removing duplicates, 518 unique individuals were retrieved as potential cases. Following individual clinical review by the corresponding reference unit, a total of 374 patients were confirmed as MND and 144 (27.8%) were excluded, most frequently because the ICD diagnostic code did not correspond to a true motor neuron disease diagnosis. MND patients were distributed across Valencia (n=196; 52.4%), Alicante (n=142; 38.0%) and Castellón (n=36; 9.6%). By phenotype, 325 were ALS (86.9%), 26 PLS 7.0%) and 23 PMA (6.1%).

### 3.2. Overall prevalence of MND and geographic distribution

The overall crude prevalence of MND on 1 January 2025 was 8.43 per 100,000 (95% CI: 7.60–9.29), and the age-adjusted prevalence was 7.85 per 100,000 (95% CI: 7.05–8.65). Prevalence was similar across the three provinces (Alicante 8.66, Valencia 8.50, Castellón 7.37 per 100,000; Table 1) and across health departments, where the wide, overlapping confidence intervals indicated no statistically significant geographical variation (Figure 1, Supplementary Table S2).

**Table 1.** Confirmed MND cases, reference population and prevalence by province, Valencian Community, 1 January 2025.

| Province | Cases | Pop. ≥15 y | Crude /100,000 (95% CI) | Adjusted /100,000 (95% CI) | Men / Women | M:F | Mean age |
| --- | --- | --- | --- | --- | --- | --- | --- |
| Alicante | 142 | 1,640,065 | 8.66 (7.26–10.12) | 8.10 (6.76–9.44) | 76 / 66 | 1.15 | 65.9 |
| Valencia | 196 | 2,305,587 | 8.50 (7.33–9.72) | 7.88 (6.77–8.99) | 123 / 73 | 1.68 | 66.2 |
| Castellón | 36 | 488,347 | 7.37 (5.12–9.83) | 6.87 (4.60–9.14) | 17 / 19 | 0.89 | 64.4 |
| **Total MND** | **374** | **4,433,999** | **8.43 (7.60–9.29)** | **7.85 (7.05–8.65)** | **216 / 158** | **1.37** | **65.9** |

*Age-adjusted rates were obtained by direct standardisation to the 2013 European Standard Population; province-level age structures were assumed to follow that of the region as a whole.*

**Figure 1.**
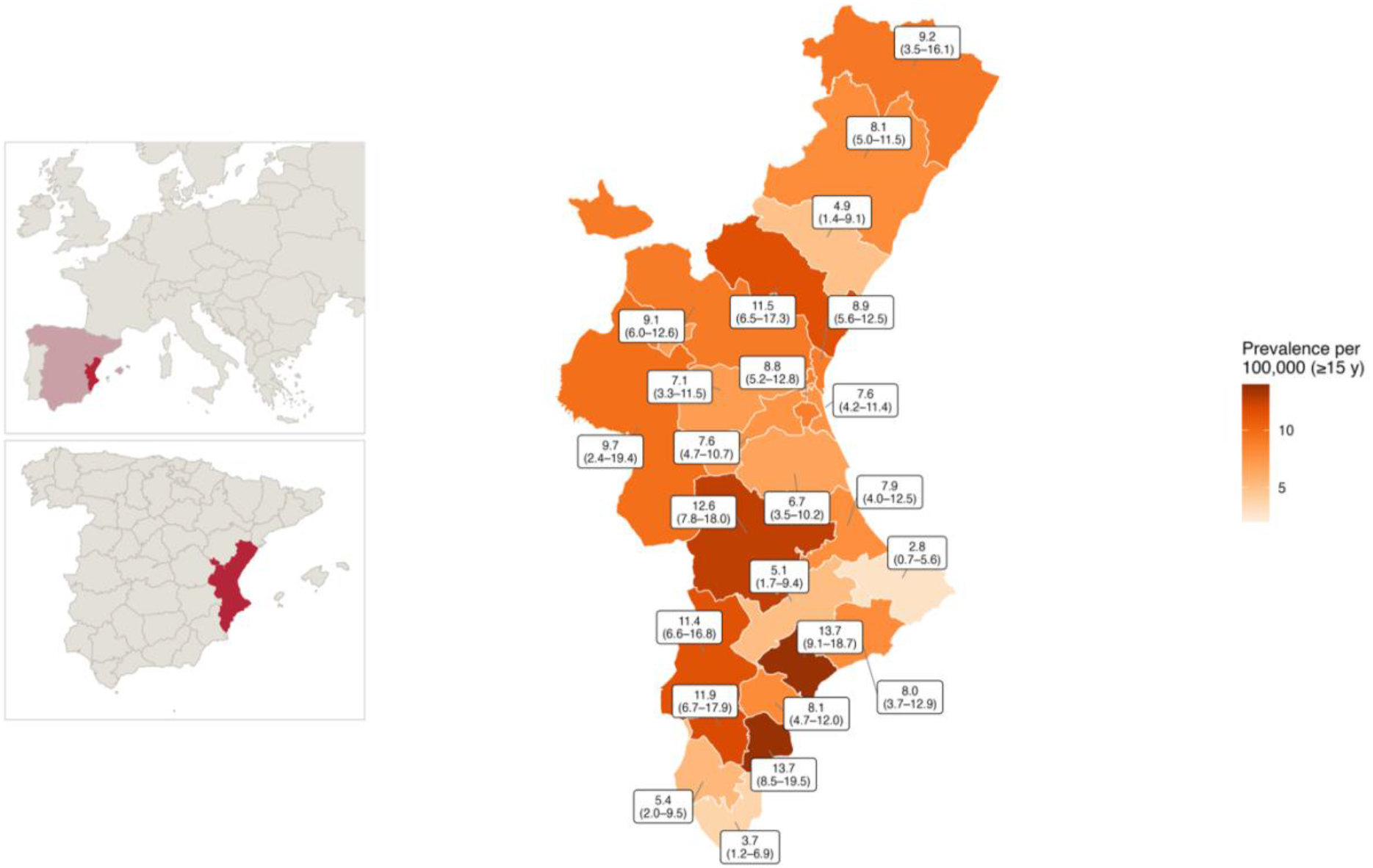
Crude prevalence of amyotrophic lateral sclerosis (ALS) per 100,000 inhabitants aged 15 years or older, by health department, Valencian Community, 1 January 2025. Each label shows the crude prevalence with its 95% Poisson confidence interval. Department names and the number of confirmed cases are provided in Supplementary Table S2. Colour intensity is proportional to crude prevalence. Confidence intervals overlap widely across departments, and the apparent geographical differences are not statistically significant, consistent with the homogeneity observed at the provincial level.

### 3.3. Prevalence by phenotype

ALS was the most frequent phenotype, with a crude prevalence of 7.33 per 100,000 (95% CI: 6.54– 8.14) and an age-adjusted prevalence of 6.84 per 100,000 (95% CI: 6.09–7.59). PLS and PMA were much rarer, with crude prevalences of 0.59 (95% CI: 0.38–0.81) and 0.52 per 100,000 (95% CI: 0.32– 0.74), respectively (Table 2).

**Table 2.**
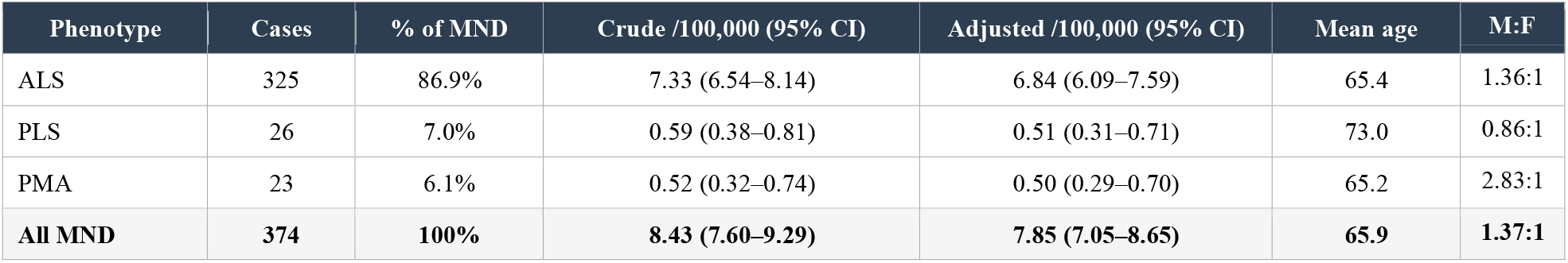
Prevalence of MND by phenotype, Valencian Community, 1 January 2025.

### 3.4. Demographic characteristics

The mean age was 65.9 years and the overall male-to-female ratio was 1.37:1. In ALS, the mean age was 65.4 years and the male-to-female ratio was 1.36:1, close to the overall figure. PMA showed a striking male predominance (2.83:1) with a similar mean age (65.2 years), whereas PLS occurred in older patients (mean age 73.0 years) and showed a slight female predominance (0.86:1). Age-specific prevalence peaked in the 70–79 year group for MND overall and for ALS, whereas the peak occurred earlier in PMA (60–69 years) and later in PLS (≥80 years), mirroring the older mean age of PLS patients (Figure 2). Riluzole was being taken by 83.4% of MND patients, with differences across phenotypes: 84.3% in ALS, 91.3% in PMA and 65.4% in PLS.

**Figure 2.**
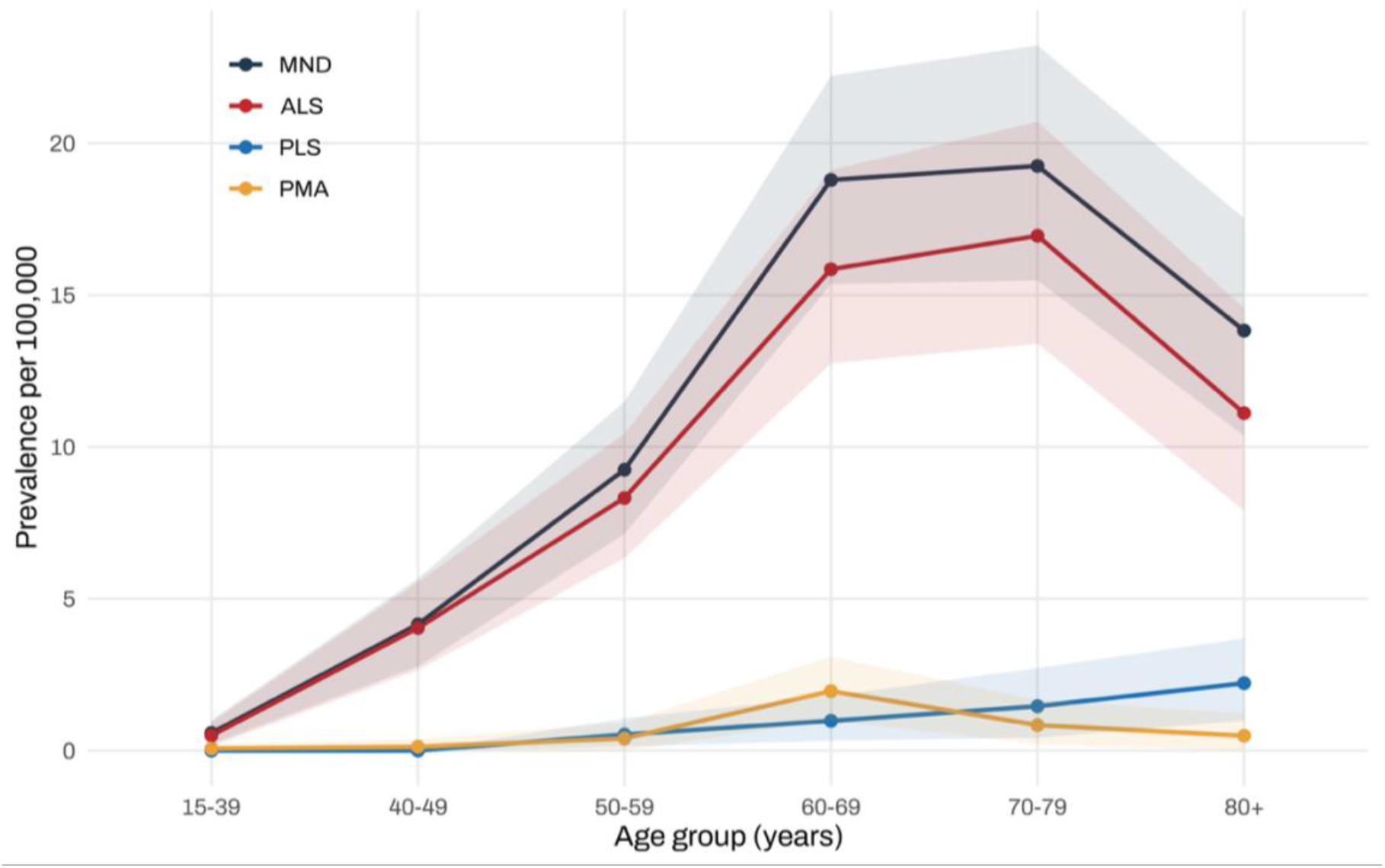
Age-specific prevalence of motor neuron disease by phenotype, Valencian Community, 1 January 2025. Crude prevalence per 100,000 population by 10-year age group for motor neuron disease overall and for each phenotype (ALS, PLS and PMA). Points represent the prevalence in each age group and shaded bands the 95% Poisson confidence intervals. Prevalence rose steeply from the fifth decade onwards and peaked in the 70–79 year group for MND overall and for ALS, whereas the peak occurred earlier in PMA (60–69 years) and later in PLS (≥80 years), mirroring the older mean age of PLS patients. The denominator for each age group was the corresponding population aged 15 years or older registered in the Primary Care Information System (SIAP).

### 3.5. Genetic findings

Genetic testing was performed in 306 of the 374 patients (81.8%); however, not all patients were tested for the same set of genes (see Methods), so the number tested differs by gene and a single denominator cannot be applied across genes. We therefore report absolute frequencies and percentages over the whole cohort (n=374), consistent with the population-based approach. A pathogenic or likely pathogenic variant was identified in 39 patients (10.4% of the cohort with a genetic yield among tested patients of 12.7%). Among these, SOD1 was the most frequent finding (17 patients; 4.5% of the cohort; 43.6% of all pathogenic variants), followed by the C9orf72 repeat expansion (13; 3.5%; 33.3% of pathogenic variants) and FUS (2; 0.5%), with single variants in ANXA11, ATP13A2, CHMP2B, SLC52A3, SQSTM1, TBK1 and VCP. Expressed as population-based estimates, the minimal prevalence of SOD1-related MND was 0.38 per 100,000 (95% CI: 0.20–0.59) and that of C9orf72-related MND was 0.29 per 100,000 (95% CI: 0.14–0.47) (Table 3, Figure 3).

**Table 3.** Genetic findings in the whole cohort (n=374), Valencian Community.

| Gene | Cases | % of cohort (n=374) | Phenotype | Population prevalence /100,000 (95% CI) |
| --- | --- | --- | --- | --- |
| *SOD1* | 17 | 4.5% | 15 ALS, 2 PMA | 0.38 (0.20–0.59) |
| *C9orf72* | 13 | 3.5% | 13 ALS | 0.29 (0.14–0.47) |
| *FUS* | 2 | 0.5% | 1 ALS, 1 PMA | 0.05 (0.00–0.11) |
| Other genes* | 7 | 1.9% | 7 ALS | 0.16 (0.05–0.29) |
| Any pathogenic or likely pathogenic variant | 39 | 10.4% | 36 ALS, 3 PMA | 0.83 (0.59–1.11) |
| No pathogenic variant identified or not tested | 335 | 89.6% | 289 ALS, 26 PLS, 20 PMA | — |

*\*ANXA11, ATP13A2, CHMP2B, SLC52A3, SQSTM1, TBK1 and VCP (one case each). Not all patients were tested for the same set of genes; percentages are calculated over the whole cohort (n=374) and, given the incomplete and heterogeneous testing, should be regarded as minimum estimates.*

**Figure 3.**
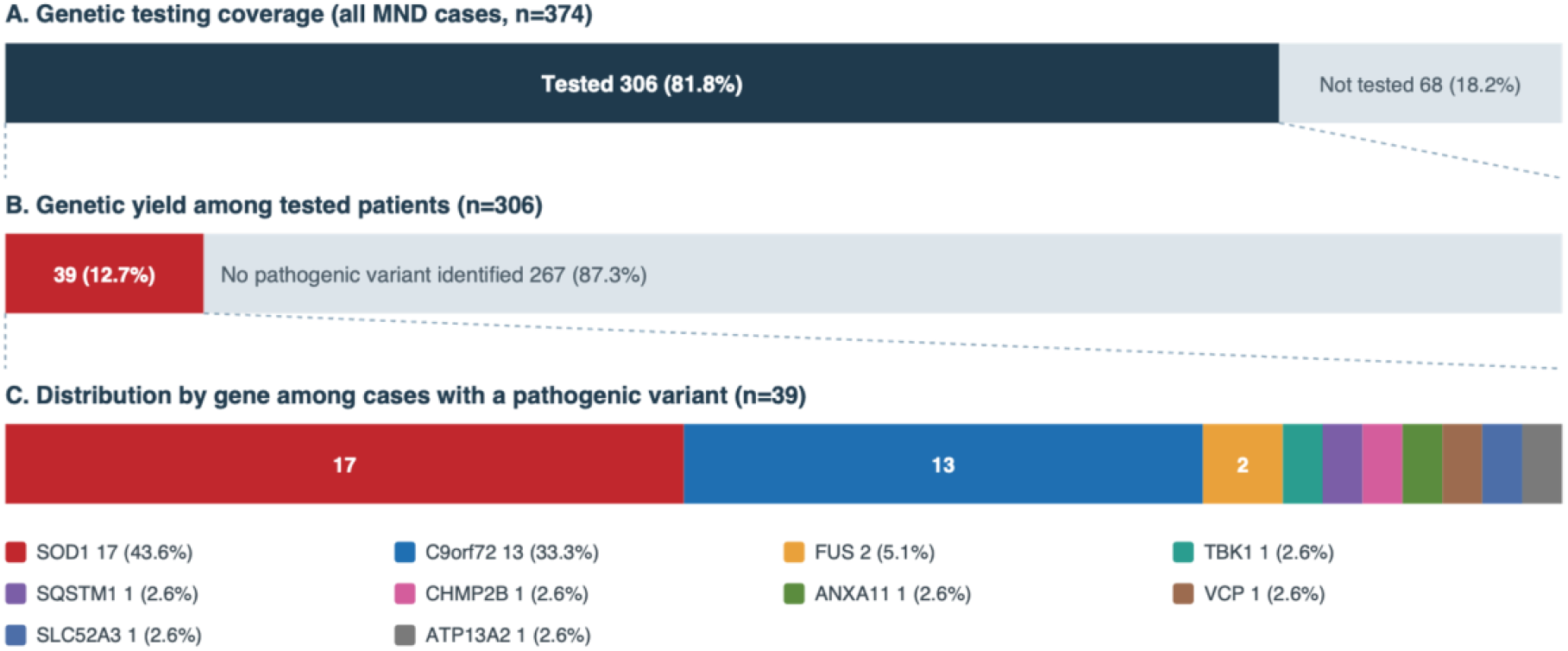
Genetic testing and findings in motor neuron disease, Valencian Community, 1 January 2025. Panel A shows the proportion of the whole cohort that underwent genetic testing. Panel B shows, among tested patients, the proportion in whom a pathogenic or likely pathogenic variant was identified. Panel C shows the distribution by gene among the 39 cases carrying a pathogenic variant. Because not all patients were tested for the same set of genes, proportions should be interpreted as estimates conditional on the testing actually performed.

## 4. Discussion

This study provides the first comprehensive, multi-source population-based estimate of MND prevalence and its subtypes in the Valencian Community. The overall MND prevalence of 8.43 per 100,000 and the ALS prevalence of 7.33 per 100,000 are consistent with the upper range of recent European estimates[3–5] and substantially higher than the 3.25 per 100,000 previously reported for Valencia in the period 2011-2019 by Barceló et al.[6]

The earlier estimate relied on a single source (reference hospital registry), whose authors cautioned that the dispersed geography of Valencia could reduce case capture[6,13]. Thus, our multi-source, four-unit approach with mandatory clinical confirmation was designed to overcome single-centre under-ascertainment, and much of the apparent increase likely reflects more complete case capture. In this study, reliance on diagnostic codes alone would have missed cases captured only through riluzole prescription, hospital admission or the units’ registries, underscoring the value of multi-source ascertainment and the necessity of individual clinical confirmation.[14,15]

A related and noteworthy finding is the proportion of administrative false positives: more than a quarter of the individuals retrieved from routine health information systems did not have motor neuron disease on clinical review,[16] most often because a G12.2 or 335.2x code had been assigned to conditions later reclassified, to suspected diagnoses never confirmed, or to coding errors.[14] This discrepancy is rarely quantified in prevalence studies, yet it has a direct methodological implication: estimates derived solely from diagnostic codes, without clinical validation, are prone to substantial overestimation. Administrative data should therefore be treated as a case-finding tool rather than a case definition, and accurate, updated clinical coding is essential both for epidemiological surveillance and for planning services for a disease with high care needs.

Beyond more complete ascertainment, several factors plausibly contribute to the higher prevalence observed. First, survival has improved: non-invasive ventilation,[17] gastrostomy[18] and structured respiratory and nutritional support are applied earlier and more widely, and a greater proportion of patients are followed in specialised multidisciplinary units, a model consistently associated with longer survival;[19–21] because prevalence is a function of both incidence and disease duration, longer survival raises prevalence even without any change in incidence. Second, progressive population ageing increases the burden of neurodegenerative disorders in general, and motor neuron disease is no exception:[22,23] our cohort included a substantial number of patients aged over 80 years, and some over 90, an age range scarcely represented in earlier series. Third, diagnostic recognition has improved, particularly among the very elderly, in whom progressive weakness was previously attributed to comorbidity, frailty or ageing itself. Thus, greater awareness among neurologists,[24] wider access to referral ALS units and the adoption of the Gold Coast criteria have facilitated case ascertainment.[25– 27] A further contributor specific to this region may be inward migration: the Valencian coast attracts residents from other regions or countries, some of whom may relocate after diagnosis, which could inflate local prevalence in particular departments. These mechanisms are not mutually exclusive and most likely act together with improved case capture, so the figures reported here are best interpreted as reflecting a combination of methodological and genuine epidemiological change.

A distinctive contribution of this study is the provision of population-based prevalence estimates for PLS and PMA, which are exceedingly rare in the literature.[10] Their frequencies (7.0% and 6.1% of MND, respectively) and their characteristic demographic profiles are consistent with previously described cohorts.[6,10,11] The homogeneity of prevalence across the three provinces, which share genetic and environmental characteristics, is best interpreted as internal validation of the ascertainment strategy rather than as true geographical variation. Population-based epidemiological data specifically addressing PMA remain extremely limited. A previous model-based study in Catalonia and Valencia estimated a PMA prevalence between 0.065 and 0.634 per 100,000 inhabitants.[6] Our crude prevalence of 0.52 per 100,000 lies within this range and provides one of the few clinically validated population-based estimates of PMA prevalence. The marked male predominance observed in PMA (male-to-female ratio 2.83:1) is consistent with the demographic profile reported in clinical lower motor neuron-predominant cohorts. [11]

In PLS sex distribution was approximately balanced, with a slight numerical female predominance (male-to-female ratio 0.86:1). Although many referral-based series have reported male predominance, recent population-based evidence does not reproduce this pattern.[6,28] A recent population-based study from the Piemonte and Valle d’Aosta ALS registry found a similar prevalence to our study (0.89 per 100,000).[28] Interestingly, they found higher age of onset than previously reported, and a male-to-female ratio of 0.58, with higher incidence in women. Our findings are consistent with these data and suggest that the marked male predominance and young age described in classical clinical PLS cohorts may partly reflect referral patterns, differences in case definition or inclusion of UMN-predominant ALS. Nevertheless, the small number of PLS cases in our cohort and the prevalence-based design preclude firm conclusions. Larger pooled population-based studies with sex-specific incidence and survival estimates are needed.

The cohort showed a high proportion of *SOD1* variants among tested patients (17 patients; 4.5% of the whole cohort; 43.6% of all pathogenic variants), exceeding the frequency (~2–3%) usually reported in Spain[29] or other countries,[30] and unexpectedly higher than the *C9orf72* expansion frequency (13 patients; 3.5% of the cohort).[30,31] This finding is of particular interest given the recent approval of the *SOD1*-targeted therapy tofersen, which makes accurate *SOD1* frequency estimates directly relevant to service planning.[32] Several considerations temper the interpretation of the genetic testing. First, genetic testing was performed in 82% of cases rather than all, and testing is preferentially offered to patients with classical ALS, a family history or younger age of onset; the denominator of tested patients is therefore enriched for a higher pre-test probability of a genetic cause, which may inflate gene frequencies relative to the whole cohort. By contrast, the *C9orf72* expansion frequency (4.2%) was within the expected range, which argues against a purely artefactual explanation and lends more weight to the *SOD1* finding. For these reasons, the population-based prevalence estimates (*SOD1* 0.38 and *C9orf72* 0.29 per 100,000), which use the whole population as the denominator, are more robust than the proportions among tested patients, and both should be regarded as minimum estimates pending complete testing. Second, while *SOD1* and *C9orf72* were tested in almost all patients, other genes were not. Thus, the frequency of other genes might be higher than reported here. Third, a regional founder effect for specific *SOD1* variants has been described in Spain and would merit dedicated haplotype analysis.[29] Fourth, most *SOD1* variants described in Spain were slowly progressive with some patients showing >20 years survival, which may further increase the prevalence of *SOD1* mutations in populations with those founder effects.[29]

The main strengths of this study are its population-based, multi-source design, the involvement of all four regional MND reference units, and the clinical validation of individual cases by experienced neurologists. This approach ensured broad geographical coverage, improved case ascertainment and allowed robust prevalence estimates not only for ALS, but also for the rarely studied PLS and PMA phenotypes.

This study has limitations. First, ascertainment depends on contact with the health system, (16) although the universal healthcare system in combination with five independent sources mitigates this. Second, some department-level denominators were estimated. Third, annual incidence could not be estimated, as the study was based on a single prevalence-day cross-section. Finally, genetic characterisation was incomplete and heterogeneous, as not all patients were tested for the same panel of genes, so the genetic frequencies reported here should be regarded as minimum estimates.

## 5. Conclusions

The crude prevalence of MND in the Valencian Community as of 1 January 2025 was 8.43 per 100,000 (age-adjusted 7.85 per 100,000), with ALS accounting for most cases (7.33 per 100,000) and PLS and PMA representing rarer subtypes. These figures are markedly higher than the previous single-unit estimate and more consistent with recent European data, underscoring the value of multi-source ascertainment. The regional network of reference units and integrated health information systems provides a robust platform for ongoing population-based MND surveillance.

## Data Availability

All data produced in the present study are available upon reasonable request to the authors

## Acknowledgements

This research was supported by the University of Alicante (Universidad de Alicante), which funded the Article Processing Charge for open access publication.

## Ethics

The study was approved by the Committee of Ethics of Clinical Research of the Dr. Balmis University Hospital (reference PI2026-018). Given the retrospective, registry-based design using anonymised data, individual informed consent was waived by the committee. The study was conducted in accordance with the Declaration of Helsinki.

## Data availability

The data that support the findings of this study are not publicly available because they contain information that could compromise the privacy of research participants and are subject to institutional and data-protection restrictions (EU Regulation 2016/679 and Spanish Organic Law 3/2018). Anonymised aggregate data are available from the corresponding author upon reasonable request.

## Declaration of generative AI and AI-assisted technologies in the manuscript preparation process

During the preparation of this work the authors used Claude (Anthropic) in order to assist with English-language editing. After using this tool, the authors reviewed and edited the content as needed and take full responsibility for the content of the published article.

## CRediT author contributions

Pablo Ros-Arlanzón: conceptualization, methodology, formal analysis, investigation, data curation, writing – original draft, writing – review and editing. Juan Francisco Vázquez-Costa: methodology, formal analysis, investigation, data curation, writing – review and editing. Ana Merchán-Cabanillas: investigation, data curation, writing – review and editing. Rosa Vilar-Ventura: investigation, data curation, writing – review and editing. Ana Pareja: investigation, data curation, writing – review and editing. Laura Lorente-Gómez: investigation, data curation, writing – review and editing. Elena Ronda-Pérez: conceptualization, methodology, formal analysis, supervision, writing – review and editing. Carmen Díaz-Marín: conceptualization, methodology, investigation, data curation, supervision, writing – review and editing.

