## Supplemental Table 1 for "Prevalence of motor neuron disease and its clinical subtypes in the Valencian Community, Spain: a population-based cross-sectional study using linked electronic health records"

**Supplementary material**

**Supplementary Table S1.** ICD codes

| **Code** | **System** | **Description** |
| --- | --- | --- |
| G12.2 | ICD-10 | Motor neuron disease |
| G12.20 | ICD-10 | Motor neuron disease, unspecified |
| G12.21 | ICD-10 | Amyotrophic lateral sclerosis |
| G12.22 | ICD-10 | Progressive bulbar palsy |
| G12.23 | ICD-10 | Primary lateral sclerosis |
| G12.24 | ICD-10 | Familial motor neuron disease |
| G12.25 | ICD-10 | Progressive spinal muscle atrophy |
| G12.29 | ICD-10 | Other motor neuron disease |
| 335.20 | ICD-9 | Amyotrophic lateral sclerosis |
| 335.21 | ICD-9 | Progressive muscular atrophy |
| 335.23 | ICD-9 | Progressive bulbar palsy |
| 335.24 | ICD-9 | Primary lateral sclerosis |
| 335.29 | ICD-9 | Other motor neuron disease |

**Supplementary Table S2** Confirmed MND cases and crude prevalence by health department, Valencian Community, 1 January 2025.

| **Code** | **Health department** | **Cases** | **Pop. ≥15 y** | **Crude /100,000 (95% CI)** |
| --- | --- | --- | --- | --- |
| 01 | Vinaròs | 8 | 86,700 | 9.23 (3.46–16.15) |
| 02 | Castelló | 22 | 260,100 | 8.46 (5.00–12.30) |
| 03 | La Plana | 7 | 143,055 | 4.89 (1.40–9.09) |
| 04 | Sagunt | 16 | 138,720 | 11.53 (6.49–17.30) |
| 05 | València–Clínic | 26 | 303,450 | 8.57 (5.60–11.86) |
| 06 | València–Arnau-Llíria | 25 | 285,657 | 8.75 (5.60–12.25) |
| 07 | València–La Fe | 22 | 250,426 | 8.79 (5.19–12.78) |
| 08 | Requena | 4 | 41,224 | 9.70 (2.43–19.41) |
| 09 | València–General | 23 | 316,911 | 7.26 (4.42–10.41) |
| 10 | València–Dr. Peset | 18 | 235,837 | 7.63 (4.24–11.45) |
| 11 | La Ribera | 15 | 225,420 | 6.65 (3.55–10.20) |
| 12 | Gandia | 12 | 151,725 | 7.91 (3.95–12.52) |
| 13 | Dénia | 4 | 143,922 | 2.78 (0.69–5.56) |
| 14 | Xàtiva–Ontinyent | 21 | 166,303 | 12.63 (7.82–18.04) |
| 15 | Alcoi | 6 | 117,045 | 5.13 (1.71–9.40) |
| 16 | La Marina Baixa | 13 | 162,996 | 7.98 (3.68–12.88) |
| 17 | Elda | 19 | 166,464 | 11.41 (6.61–16.82) |
| 18 | Sant Joan | 30 | 219,351 | 13.68 (9.12–18.69) |
| 19 | Elx | 21 | 153,459 | 13.68 (8.47–19.55) |
| 20 | Alacant | 19 | 234,090 | 8.12 (4.70–11.96) |
| 21 | Orihuela | 8 | 147,390 | 5.43 (2.04–9.50) |
| 22 | Torrevieja | 6 | 160,395 | 3.74 (1.25–6.86) |
| 23 | Manises | 13 | 182,638 | 7.12 (3.29–11.50) |
| 24 | Elx–Crevillent | 16 | 134,385 | 11.91 (6.70–17.86) |
|  | **Total** | **374** | **4,433,999** | **8.43 (7.60–9.29)** |

*Confidence intervals are wide for departments with small case counts; apparent differences between departments were not statistically significant.*
